# The effect of genetic predisposition to Alzheimer’s disease and related traits on recruitment bias in a study of cognitive ageing

**DOI:** 10.1101/2023.05.10.23289642

**Authors:** Lina M. Gomez, Brittany L. Mitchell, Kerrie McAloney, Jessica Adsett, Natalie Garden, Madeline Wood, Santiago Diaz-Torres, Luis M. Garcia-Marin, Michael Breakspear, Nicholas G. Martin, Michelle K. Lupton

**Affiliations:** QIMR Berghofer Medical Research Institute, Brisbane, QLD, Australia; School of Biomedical Sciences, Faculty of Medicine, The University of Queensland, Brisbane, QLD, Australia; Mental Health & Neuroscience Program, QIMR Berghofer Medical Research Institute, Brisbane, QLD, Australia; The University of Newcastle, NSW, Australia; School of Biomedical Sciences, Faculty of Health, Queensland University of Technology., QLD, Australia

**Author notes:** Corresponding author at: Genetic Epidemiology, QIMR Berghofer Medical Research Institute.

**Keywords:** Alzheimer’s disease, recruitment, bias, genetics, polygenic, PRS

## Abstract

The recruitment of participants for research studies may be subject to bias due to an overrepresentation of those more willing to participate voluntarily. No study has analysed the effect of genetic predisposition to Alzheimer’s disease (AD) on study participation. The Prospective Imaging Study of Ageing (PISA), aims to characterise the phenotype and natural history of healthy adult Australians at high future risk of AD. Participants approached to take part in PISA were selected from existing cohort studies with available genome-wide genetic data for both successfully and unsuccessfully recruited participants, allowing us to investigate the genetic contribution to voluntary recruitment. From a recruitment pool of 13,432 individuals (age 40-80), 64% of participants were successfully recruited into the study. Polygenic risk scores (PRS) were computed in order to test to what extent the genetic risk for AD, and related risk factors (including educational attainment, household income and IQ), predicted participation in PISA. We examined the associations between PRS and *APOE* ε4 with recruitment using logistic regression models. We found significant associations of age and sex with study participation, where older and female participants were more likely to complete the core module. We did not identify a significant association of genetic risk for AD with study participation. Nonetheless, we identified significant associations with genetic scores for key causal risk factors for AD, such as IQ, household income and years of education. Our findings highlight the importance of considering bias in key risk factors for AD in the recruitment of individuals for cohort studies.

---

The recruitment process for participants in research studies can be biased due to factors affecting their likelihood to volunteer, resulting in a sample that is not necessarily representative of the general population (Oswald et al., 2013). Recruitment bias can be exacerbated when the target population is composed of individuals with a specific disorder or related history, which may affect their willingness to participate (Patten, 2000). For instance, individuals with lower cognitive function may be more willing to enrol in research, as they may perceive themselves as being in greater need of interventions (Li et al., 2022). Similarly, people with dementia or a family history of dementia may be motivated to participate in research as a way to contribute to the development of treatment strategies (Milani et al., 2021). Several factors, such as lack of confidence, depressive symptoms (Ghanbari et al., 2023), unemployment and low educational attainment (Haring et al., 2009) may lead to a decline in participation while willingness to participate has been shown to be associated with favourable profiles, such as younger age and higher education attainment (Ganguli et al., 2015), (Enzenbach et al., 2019).

Recruitment bias can impact prevalence estimates. In addition, it can affect association estimates, caused by collider bias, where two variables independently influence a third collider variable. Conditioning on the collider (in this case, study participation) can contribute to a spurious association between the two variables (Zheng et al., 2012).

Alzheimer’s disease (AD) is a weakening neurological condition, characterized by progressive neurodegeneration and decline of cognitive function leading to dementia (Andrews et al., 2021). The symptomatic burden of dementia typically occurs late in life, but it is preceded by a long preclinical phase, characterised by neuropathological impairment. Therefore, much of the ongoing dementia research is focused on elucidating early pathological changes (Jack et al., 2018). However, no study has analysed the effect of genetic predisposition to AD and causal traits on study participation in middle aged and older individuals.

The Prospective Imaging Study of Ageing: Genes, Brain and Behaviour (PISA), aims to characterise the phenotype and natural history of healthy adult Australians at high future risk of Alzheimer’s disease (Lupton et al., 2021). Potential PISA participants were selected from previous genetic cohort studies on the condition that they had genome-wide genetic data already available. This offers a unique opportunity to investigate the genetic contribution to voluntary recruitment, enabling us to test for differences between those successfully recruited into PISA and those who were not.

In the present study, we aimed to investigate whether the likelihood of participating in the PISA study was influenced by factors associated with a higher risk of AD, including genetic risk of AD and causally associated risk factors. Using Mendelian randomisation, we and others have previously shown that out of several potentially modifiable AD risk factors educational attainment (EA), intelligence (measured by intelligence quotient (IQ)) and socioeconomic status (measured by household income (HI)) have a causal association with AD (Thorp et al., 2022). Therefore, we used polygenic risk scores (PRS) to assess the relationship between the genetic risk for AD and educational attainment, intelligence, and household income. By examining the association between these factors and the likelihood of participating in the PISA study, we aimed to assess whether there is a heritable component that is mediating agreement to participate in PISA, and if this could create bias in the investigation of risk factors and prodromal disease markers for AD.

## Materials and Methods

### Sample recruitment

PISA is a prospective cohort study of healthy Australians in mid to late adulthood. The sample recruitment pool consisted of 15,531 participants with available genome wide genotyping, derived from extensive in-house cohorts, including studies of Australian twins and their families, and genome wide association studies (GWAS) for physical or psychiatric conditions conducted over four decades at QIMR Berghofer (Heath et al., 2011; Medland et al., 2009). We attempted to re-contact all research participants between 40 to 80 years old (years of birth: 1946–1986). Those who were known to be deceased, were living overseas, had requested not be included in further studies, or had no available valid contact details were excluded, resulting in a final sample size of 13,432. Over nine thousand participant records (N=9,685, 72%) were successfully updated. The majority (61.7%) of the participants in the study were women with a mean age of 58.8 and standard deviation of 7.4 years. All re-contacted participants were invited to complete an online survey that aimed to update aspects related to lifestyle and cognitive and behavioural function (full details are given in (Lupton et al., 2021)). The survey began with an information and consent page followed by a core module that captured the central dataset. Once the core module was complete, the survey included ten additional modules and the option to consent to linking medical records (described in full in (Lupton et al., 2021)). No incentive or remuneration was offered to participants to take part in this stage of the study. Participants were offered the option of completing a paper version of the survey, which was mailed to them upon request. Subjects were informed that they might be invited to take part in further research. Overall, 4,801 participants completed the core module of the online PISA survey, with a mean age of 60 years and standard deviation of 7 years.

For the purposes of this study, those who took part by completing the core survey module of the PISA questionnaire were considered as participating. All participants provided online informed consent prior to participating in the study, which included permission to link to previously collected data. The PISA study protocol (P2210) was approved by the Human Research Ethics Committees (HREC) of QIMR Berghofer Medical Research Institute.

### SNP-based heritability

Genome-wide genotyping of participants within our recruitment pool was performed using a range of genotyping arrays as previously detailed (Cuellar-Partida et al., 2015; Medland et al., 2009). Datasets were combined with strict quality control procedures and imputed to the Haplotype Reference Consortium (HRC) Release 1 reference panel (McCarthy et al., 2016).

We employed genome-based restricted maximum likelihood (GREML) as implemented in GCTAv.1.91.3beta (Yang et al., 2011) to estimate the proportion of variance in the PISA study (on the observed scale) explained by measured genetic differences (SNP-based heritability). This approach leverages a genetic-relatedness matrix (GRM) and restricted maximum likelihood to partition the variance of a phenotype into genetic and environmental components (Yang et al., 2010). This method is frequently used to assess whether genetic covariation explains a significant proportion of the covariation of a trait. For this study, a GRM based on a subset of unrelated individuals of European ancestry was leveraged to identify evidence for a genetic component to taking part in PISA.

### Polygenic risk scores (PRS)

A PRS is a way to estimate an individual’s genetic susceptibility for a particular trait or disease based on their genetic makeup. It is calculated using the sum of an individual’s genome-wide or location specific genotypes, weighted by effect size estimates for single nucleotide polymorphisms (SNPs) associated with the trait or disease of interest (Clark et al., 2022).

We computed PRS, using the SBayesR approach, to investigate the extent to which the genetic risk for AD, EA, IQ, and HI predicted participation in PISA. We estimated PRS using GWAS summary statistics for AD (Bellenguez et al., 2022), EA (Lee et al., 2018), IQ (Savage et al., 2018) and HI (Hill et al., 2019). For the AD PRS, SNPs within 500kb distance at either side of the *APOE* locus were excluded to ensure the exclusion of the *APOE*-associated signal. *APOE* genotype was considered separately from the AD PRS (derived from SNPs rs429358 and rs7412) and was either obtained from genome-wide SNP chip data (where either the *APOE* SNPs were directly genotyped on the array or imputed with a high degree of certainty) or directly genotyped using TaqMan SNP genotyping assays, as described in (Lupton et al., 2018). Furthermore, as both the GWAS for EA and IQ included potentially overlapping samples with the PISA cohort, we computed PRS using summary data from GWAS meta-analyses that exclude QIMR Berghofer cohorts, including any PISA participants and their family members (referred to as ‘leave one out’ (LOO) QIMRB summary statistics). Prior to estimating SBayesR PRS, we excluded low imputation quality (r^2^□<□0.6), allele frequency (MAF < 0.01), non-autosomal, and strand-ambiguous variants. Imputed genotype dosage data were used to calculate PRS by multiplying the variant effect size by the dosage of the effect allele. Finally, the total sum was calculated across all variants. The SBayesR PRS was generated using Plink 2.0 (Chang et al., 2015; Purcell et al., 2007).

### Statistical analyses

We examined the association of PRS for AD, EA, IQ and HI PRS and also *APOE* ε4 carrier status with recruitment bias using a logistic regression model in GCTA (1.91.3beta), which accounted for sex, age, and the first five genetic ancestry principal components (PCs) as fixed effects. Relatedness among individuals was accounted for as a random effect with a genetic relatedness matrix. The AD PRS excludes the *APOE* region and is therefore independent of *APOE* ε4. A Nagelkerke’s R^2^ was used to estimate the variance explained by the PRS or *APOE* status.

## Results

### Demographic factors

Participants who took part in PISA were, on average, older than those who did not (OR=1.007; 95%C.I.=[1.006-1.008] per year of age) *p-value=7*.*33E-30 (Table 1)*. Considering the participation rate stratified by decade showed that the largest number of participants were in the 50-69 age range, representing 78.6% of the whole sample (*Table 2*). Female participants were more likely to participate in PISA than males *(Table 1)* (OR=1.063; 95%I.C.=[1.04-1.08]) *p-value=1*.*18E-12*.

**Table 1.** Demographics of participants to take part in PISA.

| Table 1. Demographics of participants to take part in PISA |  |  |  |  |
| --- | --- | --- | --- | --- |
|  | Whole sample<br>(n=13432) | Participated in PISA<br>(n=4801) | Not<br>Participated in<br>PISA<br>(n=8631) | Participated in<br>PISA OR (95% C.I) |
| Age in years |  |  |  |  |
| Mean<br>(SD) | 59.1 (7.4) | 60.08 (7.15) | 58.8 (7.4) | 1.03 (1.03 - 1.04) |
| Female | 8,548 (63.6%) | 3,219 (67%) | 5,329(61.7%) | 1.31 (1.23 - 1.39) |
| Male | 4,884(36.4%) | 1,582 (33%) | 3,302 (38.3%) |  |
| APOE<br>□4 +ve | 3,935 (29.3%) | 1,411 (32.2%) | 2,524 (31.35%) | 1.02 (0.94 - 1.09) |
| APOE<br>□4 -ve | 9,206(70.7%) | 3,281 (67.8%) | 5,925 (68.6%) |  |

(291 Missing data proportion of APOE )

**Table 2.** Participation rate by decade.

(291 Missing data proportion of APOE )
| **Table 2. Participation rate by decade** |  |  |
| --- | --- | --- |
| **Age-group** | **N** | **% participated in PISA in**<br>**each age group** |
| **< 40** | 1 | 0.01 |
| **40-49** | 1567 | 11.67 |
| **50-59** | 5907 | 43.97 |
| **60-69** | 4789 | 35.65 |
| **70-80** | 1151 | 8.57 |
| **> 80** | 17 | 0.13 |

### Genetic Factors

The GREML analysis suggested the presence of genetic contribution to the likelihood of participating in PISA. The SNP-based heritability on the observed scale was 0.14 (SE = 0.013, *p= <0*.*0001*, phenotypic variance of the observed scale ∼0.22).

The results of the association of genetic variables (PRS and APOE), sex, and age with participation in the PISA study are shown in Table 3 and Figure 1. Neither AD PRS nor presence of *APOE* □4 were associated with participation in PISA. The EA, IQ and HI PRS were positively associated with participating in PISA.

**Table 3.**
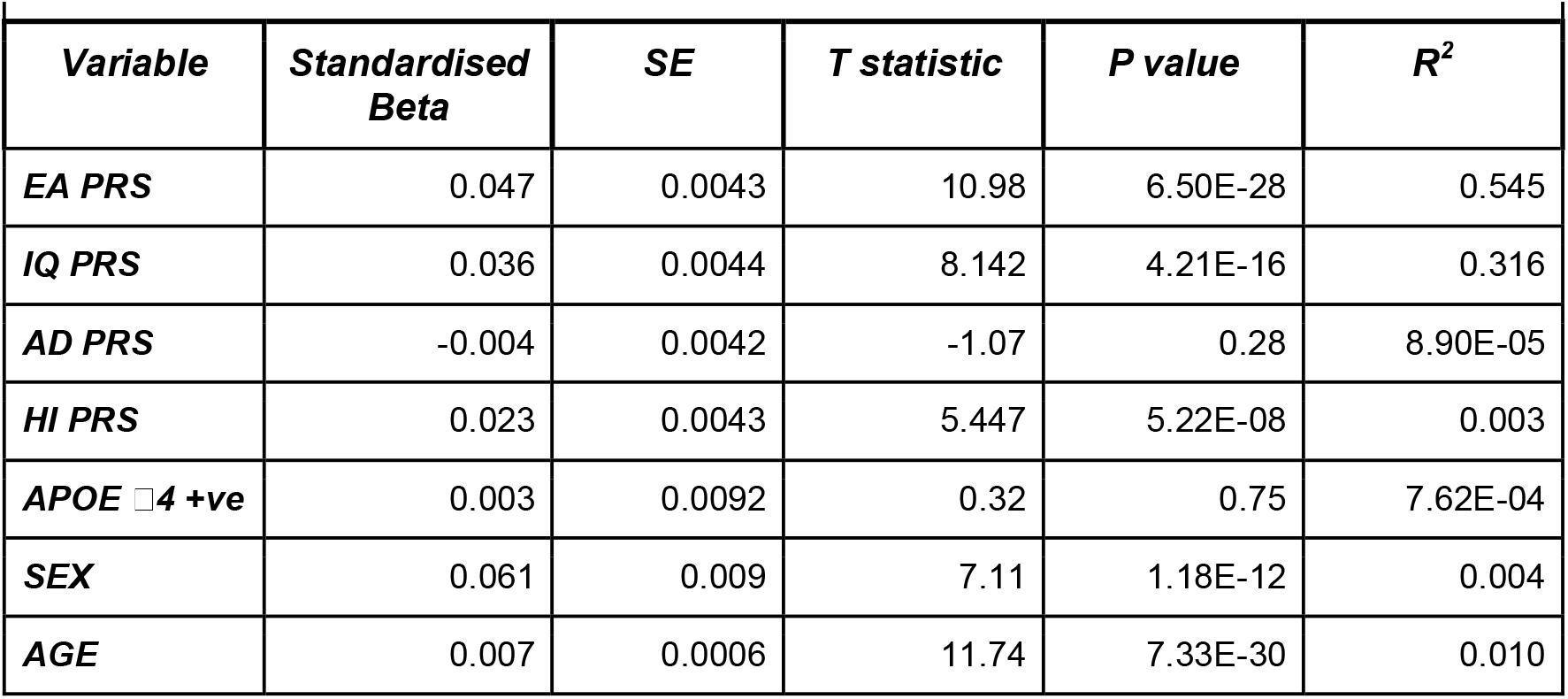
The Association of genetic variables, sex, and age with participation in the PISA study, p < 0.0083 (bonferroni corrected threshold).

| **Variable** | **Standardised Beta** | **SE** | **T statistic** | **P value** | **R<sup>2</sup>** |
| --- | --- | --- | --- | --- | --- |
| **EA PRS** | 0.047 | 0.0043 | 10.98 | 6.50E-28 | 0.545 |
| **IQ PRS** | 0.036 | 0.0044 | 8.142 | 4.21E-16 | 0.316 |
| **AD PRS** | -0.004 | 0.0042 | -1.07 | 0.28 | 8.90E-05 |
| **HI PRS** | 0.023 | 0.0043 | 5.447 | 5.22E-08 | 0.003 |
| **APOE ε4 +ve** | 0.003 | 0.0092 | 0.32 | 0.75 | 7.62E-04 |
| **SEX** | 0.061 | 0.009 | 7.11 | 1.18E-12 | 0.004 |
| **AGE** | 0.007 | 0.0006 | 11.74 | 7.33E-30 | 0.010 |

**Fig 1.**
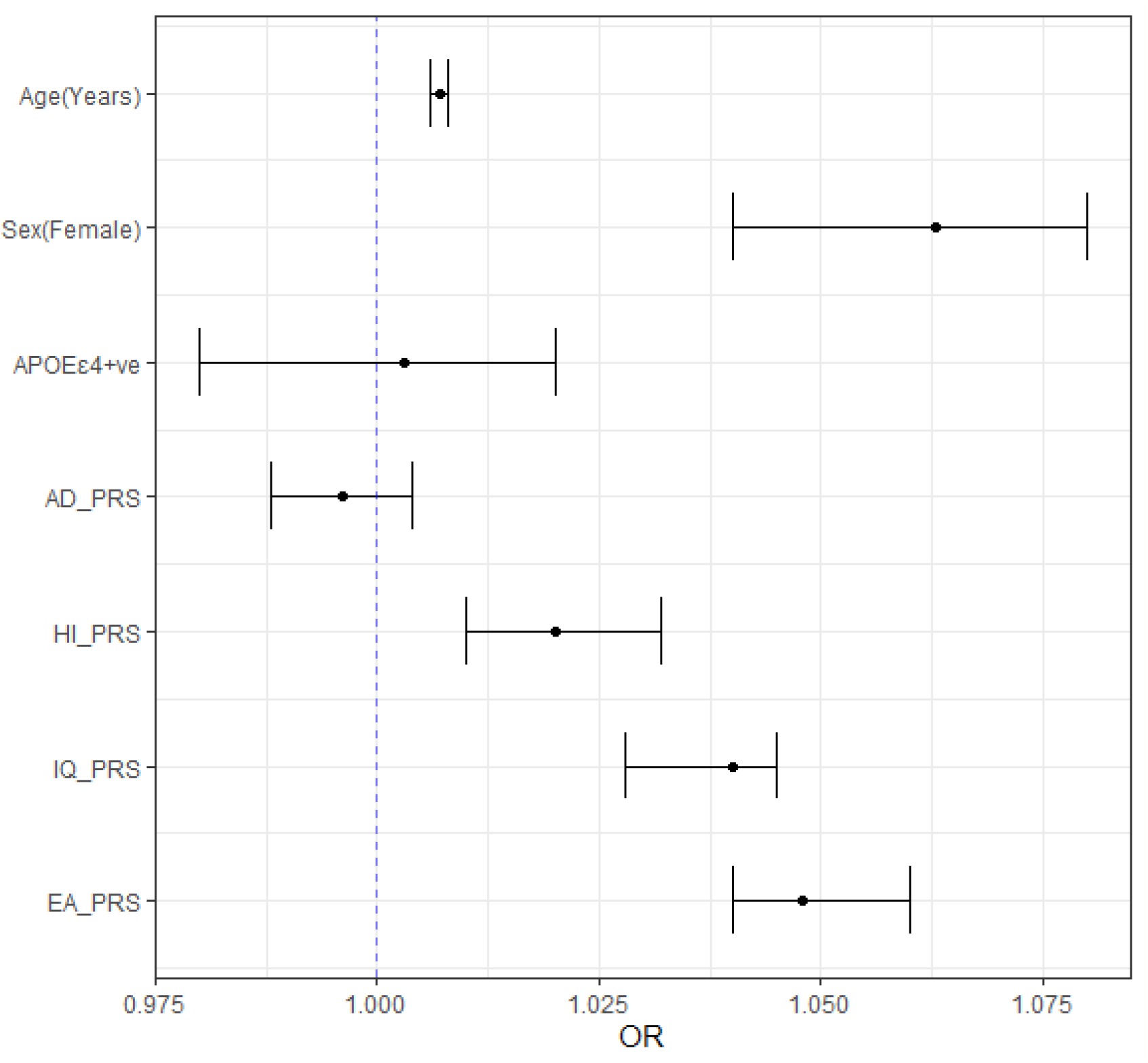
Forest plots depict odds ratios (OR) and 95% confidence intervals for the association between age, sex, PRS and presence of APOE □4 with participation in the PISA study.

## Discussion

We investigated systematic differences between individuals who decided to participate in the PISA study (i.e., complete the core survey module) with those who, despite being contacted and participating in previous studies, did not participate. Investigating variables that associate with study participation is invariably challenging, since data are required to be collected on individuals who are not willing to participate in a study. The PISA study offers a unique opportunity in this research area, as the recruitment pool consists of participants from previous studies, where genetic data has already been collected. PISA consists of middle aged and older individuals aged ≥ 40 yrs with an aim of discovering markers of early Alzheimer’s disease pathology to identify modifiable risk factors, and establish the very earliest phenotypic and neuronal signs of disease onset. Therefore, it is crucial to identify factors that are associated with the likelihood of recruitment into the study and have the potential to introduce bias in association estimates in downstream analyses.

Analysis of the relationship between sex, age and study participation revealed that older female individuals were more likely to participate in PISA (Tables 1 and 2, Figure 1). It is well known that women are more likely than men to participate in clinical research. Research into the factors that influence women’s decision to participate has found that decisions may be impacted not only by individual characteristics, but also by interpersonal relationships and social norms within the community (Baker et al., 2005; Liu & Dipietro Mager, 2016). We found in our recruitment pool of middle aged and older individuals that participants aged 50 and older were more likely to participate than those in their 40s, which may reflect a lack of spare time in the younger age group who are more likely to be of working age and with younger families. It is also possible that older participants perceived more personal risk of cognitive decline and dementia. The recruitment rate reduces once participants reach 70. These finding echoes those in studies of age related diseases, where the odds of participation are greater in older participants (aged around 55-75 years) and reduced in younger age groups, and once participants reach over 75 years (*American Journal of Epidemiology*, n.d.; Beard et al., 1994; Jacobsen et al., 2004).

Using a PRS approach we tested whether the genetic liabilities to AD and causally associated traits are associated with the decision to participate in the PISA study. We tested for an association of EA, IQ and HI, which are highly related to each other with complex multifaceted relationships. We have previously shown all three to be causally associated with risk of AD at a genetic level using mendelian randomisation analysis, and that this causal association is led by the cognitive component of EA (intelligence). (Thorp et al., 2022).

We did not find evidence suggesting that the genetic risk of AD, for both AD PRS (with *APOE* region removed) or *APOE* genotype, are associated with recruitment bias in our sample of middle aged and older research participants. However, we identified an association of PRS for key causal risk factors for AD, EA, IQ and HI. Previous studies have highlighted the strong relationship between education level and voluntary participation (Enzenbach et al., 2019; Lissner et al., 2003; Van Loon et al., 2003). The over representation of more highly educated individuals in epidemiological studies is likely due to greater health awareness and interest in science (Galea & Tracy, 2007). In addition, the association of low socioeconomic status with reduced participation in cohort studies and clinical trials has been well documented, and is thought to be due to several factors, including psychosocial and practical barriers to participation.(Howe et al., 2013; Stuber et al., 2020).

A recent investigation of genomic predictors of participation in optional components of the large scale UK Biobank study (up to 451,306 participants) also identified an association of genetic predictors of educational duration, intelligence and lower levels of deprivation (using four measures) with participation, as well as a negative correlation with AD in this well powered sample. Causal associations were confirmed using Mendelian randomisation (Tyrrell et al., 2021). We did not replicate these previous findings of an association of recruitment with genetic risk for AD including an association with APOE e4 carrier status (Tyrrell et al., 2021). This may be due to decreased power in our dataset, or potentially the influence of an opposing bias where those with a strong family history of AD due to inheritance of *APOE* 4 are more likely to be compelled to volunteer for a study researching ageing and dementia.

Limitations of the present study must be acknowledged. The participants in the recruitment pool had all previously been successfully recruited into genetic studies, and are therefore more likely to volunteer again. These baseline studies are not representative of the current Australian population, comprising predominantly those of White European ancestry, an over-representation of females, and individuals recruited based on a phenotypes of interest including twin pairs and their family members, as well as physical and psychiatric conditions. In this study, a lack of response was interpreted as a passive refusal to participate, but a subset of the recruitment pool may have out of date contact details. This was mitigated as much as possible by updating contact details using electoral roll information, contacting participants through several modalities including email, letter and phone when available, and using available death records to exclude those who had died. As much as possible we aimed to prevent accessibility issues for any participants without internet access or computing capabilities by making paper copies of the survey available upon request.

Overall this work shows that in the PISA study, key causal risk factors for Alzheimer’s disease related to cognition are associated with recruitment. Therefore, participants are not accurately representative of the population. Although this does not necessarily invalidate study findings in the investigation of risk factors for age related cognitive decline and dementia, it must be considered when interpreting findings, specifically for informing analysis strategies, using the appropriate control variables and interpreting the generalizability of findings to the general population (Elwood, 2013; Rothman et al., 2013).

## Data Availability

All data produced in the present study are available upon reasonable request to the authors

## Acknowledgements

We would like to acknowledge the contribution of the research participants. The population based sample for PISA comprises of research participants who have taken part in genetic epidemiology studies led by NM (with colleagues and collaborators) at QIMR Berghofer and elsewhere over the past 40 years. Demographic and contact information as well as genome-wide SNP chip data were utilised from these previous studies for participant selection and recruitment. Principal sources of funding for these early studies were from grants to NGM from Australian NHMRC and to Andrew Heath and Pam Madden (Washington University, St Louis) from NIH (mainly NIAAA and NIDA) and we gratefully acknowledge these contributions.

## Ethical standards

The authors assert that all procedures contributing to this work comply with the ethical standards of the relevant national and institutional committees on human experimentation and with the Helsinki Declaration of 1975, as revised in 2008.

